# Training parameters and longitudinal adaptations that most strongly mediate walking capacity gains from high-intensity interval training post-stroke

**DOI:** 10.1101/2023.02.20.23286194

**Authors:** Pierce Boyne, Allison Miller, Sarah M. Schwab, Heidi Sucharew, Daniel Carl, Sandra A. Billinger, Darcy S. Reisman

## Abstract

**Background:** Locomotor high-intensity interval training (HIIT) has been shown to improve walking capacity more than moderate-intensity aerobic training (MAT) after stroke, but it is unclear which training parameter(s) should be prioritized (e.g. speed, heart rate, blood lactate, step count) and to what extent walking capacity gains are the result of neuromotor versus cardiorespiratory adaptations.

**Objective:** Assess which training parameters and longitudinal adaptations most strongly mediate 6-minute walk distance (6MWD) gains from post-stroke HIIT.

**Methods:** The HIT-Stroke Trial randomized 55 persons with chronic stroke and persistent walking limitations to HIIT or MAT and collected detailed training data. Blinded outcomes included 6MWD, plus measures of neuromotor gait function (e.g. fastest 10-meter gait speed) and aerobic capacity (e.g. ventilatory threshold). This ancillary analysis used structural equation models to compare mediating effects of different training parameters and longitudinal adaptations on 6MWD.

**Results:** Net gains in 6MWD from HIIT versus MAT were primarily mediated by faster training speeds and longitudinal adaptations in neuromotor gait function. Training step count was also positively associated with 6MWD gains, but was lower with HIIT versus MAT, which decreased the net 6MWD gain. HIIT generated higher training heart rate and lactate than MAT, but aerobic capacity gains were similar between groups, and 6MWD changes were not associated with training heart rate, training lactate, or aerobic adaptations.

**Conclusions:** To increase walking capacity with post-stroke HIIT, training speed and step count appear to be the most important parameters to prioritize.

## INTRODUCTION

Locomotor high-intensity interval training (HIIT) involves bursts of fast walking alternated with recovery periods, and is a promising strategy for recovering walking capacity (i.e. speed and endurance) after stroke.^1-6^ For example, the multi-center HIT-Stroke Trial recently found that locomotor HIIT improved 6-minute walk distance (6MWD) and other outcomes significantly more than conventional moderate-intensity aerobic training (MAT).^4^ Still, HIIT protocols have varied widely in previous studies,^e.g.4-6^ and the optimal protocol for maximizing walking recovery remains uncertain.

When designing a HIIT protocol, burst and recovery duration can be manipulated to prioritize different training parameters (e.g. training speed, heart rate, blood lactate, or step count).^1,7,8^ For example, training speed can be prioritized by using shorter burst durations^7^ and/or longer recovery periods.^8^ However, it remains unclear which training parameters are the most important to prioritize. This would be difficult to test using standard clinical trial analysis methods because it would require a very large number of treatment groups to compare all the possible training parameter combinations.

Mediation analysis may be able to address this problem, by assessing which training parameters lead to (i.e. *mediate*) the greatest improvements in walking capacity. This analysis approach is powerful for evaluating the processes by which interventions lead to outcomes (i.e. how an intervention works or its mechanisms of action), especially in the context of a randomized trial.^9,10^ Figure 1 illustrates the mediation analysis concept and shows a simplified example testing how much of the net difference in 6MWD gains between HIIT and MAT is attributable to a particular hypothesized mediator (e.g. training speed). The ‘a’ effect quantifies the extent to which these treatment protocols differentially engage the mediator, while the ‘b’ effect estimates the influence of that mediator on the outcome. Thus, the overall ‘a x b’ mediation effect quantifies the extent to which these treatment protocols differentially affect the outcome through the mediator.

**Figure 1.**
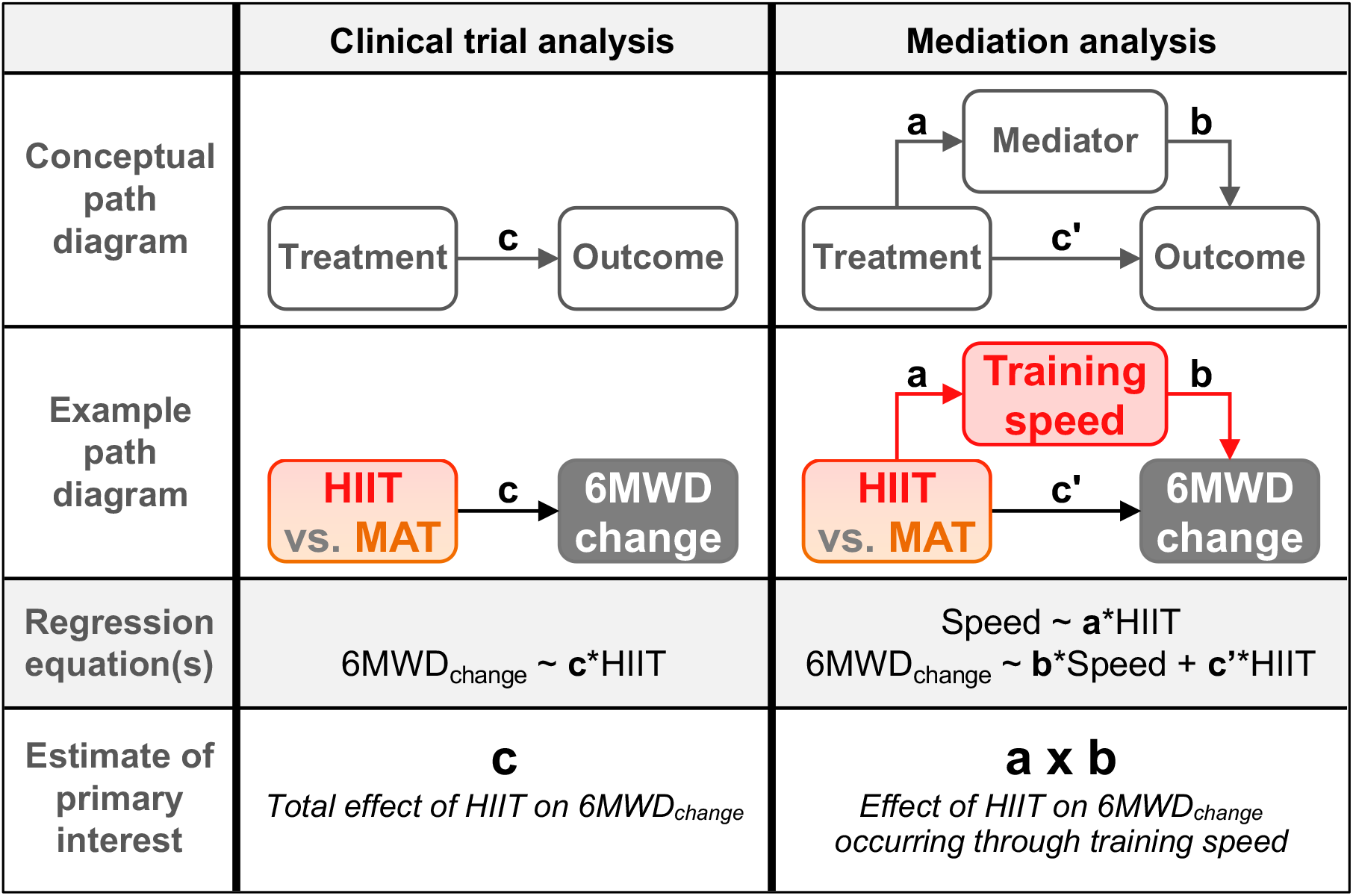
Mediation analysis concept illustration. The simplest possible analyses are shown for the effects of high-intensity interval training (HIIT) versus moderate intensity aerobic training (MAT) on 6-minute walk distance (6MWD). A standard clinical trial analysis would estimate **c**, the total net effect of HIIT on 6MWD change. Mediation analysis estimates **a**, the net effect of HIIT on a hypothesized mediator (e.g. training speed); **b**, the effect of the mediator on 6MWD change (while controlling for treatment group); and **c’**, any residual net effect of HIIT on 6MWD change after accounting for the mediator. The estimate of primary interest is the product of **a** x **b**, which is known as the indirect path or mediated effect. This represents the net effect of HIIT on 6MWD change occurring through the mediator. In the mediation model, the total net effect of HIIT on 6MWD change can be calculated as the sum of the indirect and residual direct paths, i.e. (**a** x **b**) + **c’**. Thus, mediation analysis can be viewed as decomposing the total effect into its mediated component and any residual effects that cannot be accounted for by the mediator variable(s) in the model.

Importantly, the ‘a’ effect and ‘a x b’ effect are protocol-specific, whereas the ‘b’ effect is controlled for treatment protocol, and is designed to provide broader insights about important mediators to target with *any* similar intervention in this population.^11,12^ Therefore, this type of analysis can evaluate which training parameters are the strongest mediators of between-group differences for these HIIT and MAT protocols (the ‘a x b’ effect), while also helping us understand which training parameters are the most important to prioritize when selecting a locomotor training protocol in general (the ‘b’ effect).

Different training parameters might also lead to walking capacity gains through different longitudinal adaptations, and this could affect which individuals are most responsive to the treatment.^13^ Thus, a second question to consider is which longitudinal adaptations mediate the greatest improvement in walking capacity. Locomotor HIIT is thought to improve walking capacity via both neuromotor gait adaptations and aerobic conditioning.^1,14^ It is plausible that HIIT protocols prioritizing training speed or stepping repetition might preferentially engage neuromotor adaptations, while protocols prioritizing heart rate might preferentially drive aerobic conditioning.^7,8^ However, no previous studies have tested the extent to which walking capacity gains after stroke are mediated through increases in neuromotor gait function versus aerobic conditioning.

The purpose of this study was to assess which specific training parameters and longitudinal adaptations most strongly mediate net gains in walking capacity (6MWD) from locomotor HIIT versus MAT post-stroke. Since the HIIT protocol involved short bursts at maximum speed and short recovery periods, we hypothesized that the net walking capacity gains from HIIT versus MAT would be significantly mediated by differences in training speed, heart rate and blood lactate (Figure 2, left panel), and by longitudinal improvements in both neuromotor gait function and aerobic capacity (Figure 3, left panel).

**Figure 2.**
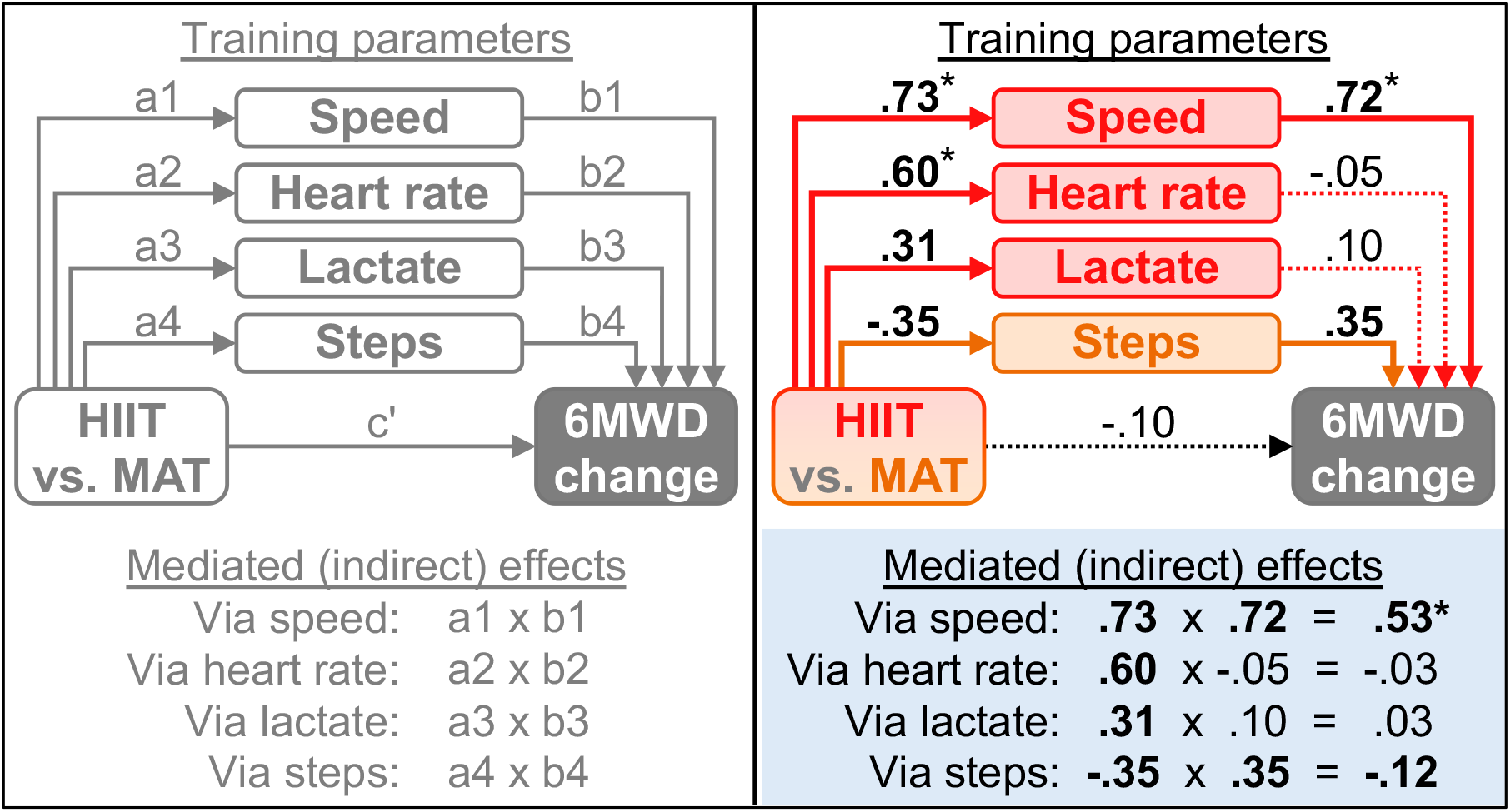
Training parameters mediating net 6MWD changes induced by HIIT. <u>Left</u>: Path diagram of structural equation model. All paths are adjusted for baseline 6MWD (not shown), baseline Fugl-Meyer lower limb motor score (not shown) and all other paths leading to the same variable. Labels for each path correspond with regression equations shown in methods. <u>Right</u>: Values are standardized path coefficients. Bolded values and solid lines are p<.05. For paths starting from the HIIT vs. MAT box, positive values indicate the training parameter was higher in the HIIT group and negative values mean it was higher in the MAT group. *Training protocol (HIIT vs. MAT) had a greater effect on training speed (treadmill and overground combined) than on all other training parameters (p≤.0002) except heart rate (p=.06), and had a greater effect on heart rate than on lactate or step count (p≤.03). Speed had a greater effect on 6MWD change than other training parameters (p≤.002) except step count (p=.14). Speed was a greater overall mediator than all other training parameters (p≤.006). After accounting for the training parameters, the remaining direct effect of training protocol on 6MWD change (c’) was not different from zero (p=0.49). HIIT, high-intensity interval training; MAT, moderate-intensity aerobic training; 6MWD, 6-minute walk distance

**Figure 3.**
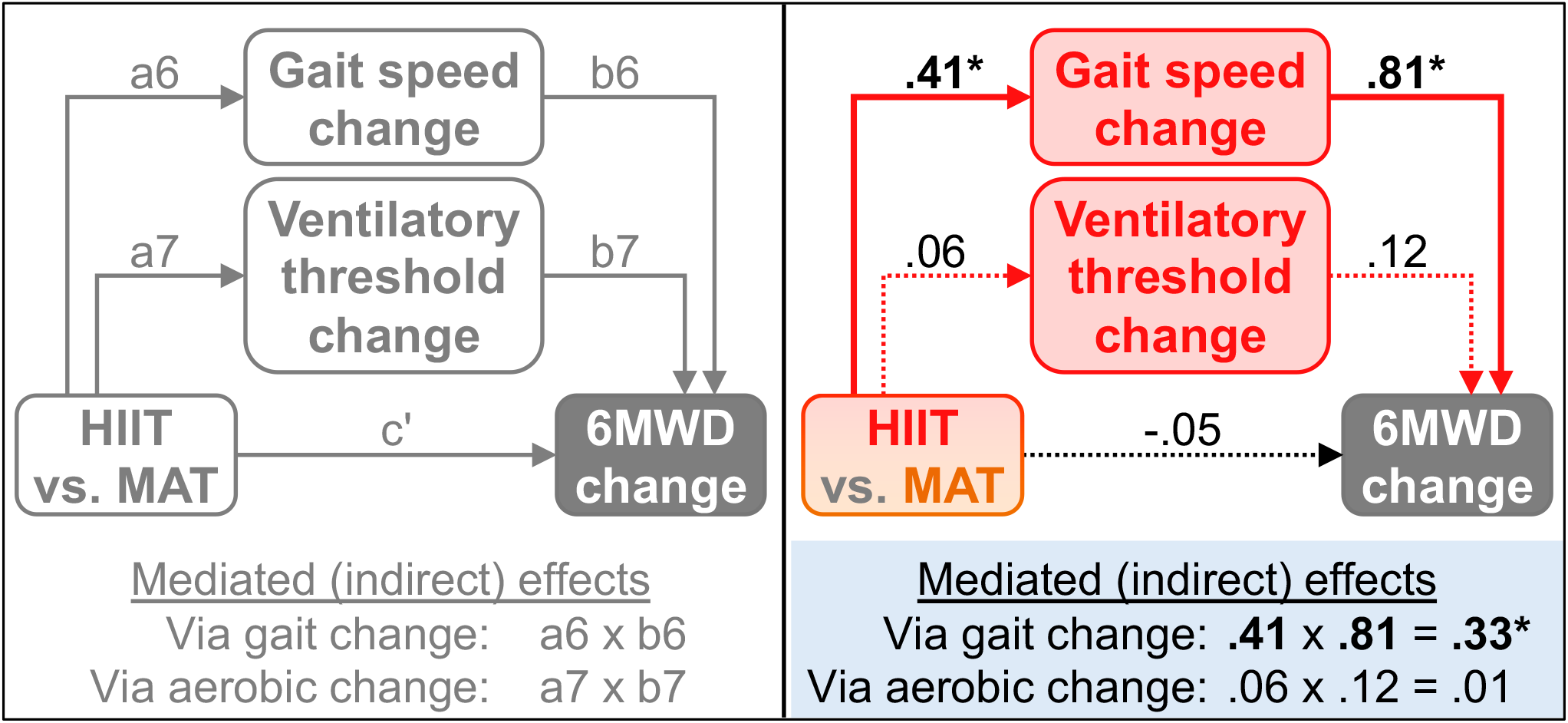
Mediating effects of longitudinal adaptations in neuromotor gait function versus aerobic capacity on net 6MWD changes induced by HIIT. <u>Left</u>: Path diagram of structural equation model. All paths are adjusted for baseline 6MWD (not shown), baseline Fugl-Meyer lower limb motor score (not shown) and all other paths leading to the same variable. Labels for each path correspond with regression equations shown in methods. <u>Right</u>: Values are standardized path coefficients. Bolded values and solid lines are p<.05. *Training protocol (HIIT vs. MAT) had a greater effect on fastest gait speed vs. ventilatory threshold (p=.01). Fastest gait speed change had a greater effect on 6MWD change than ventilatory threshold change (p<.01) and was a greater overall mediator (p<.01). HIIT, high-intensity interval training; MAT, moderate-intensity aerobic training; 6MWD, 6-minute walk distance

## METHODS

This was an ancillary analysis using data from the HIT-Stroke Trial, which has been reported in detail elsewhere (https://clinicaltrials.gov/ct2/show/NCT03760016).^4,14^ Briefly, 55 ambulatory participants >6 months post-stroke with persistent walking limitations (self-selected gait speed <1.0 m/s) and stable condition were randomized to either HIIT or MAT; each involving 45 minutes of walking exercise, 3 times/week for 12 weeks. Each session included a 3-minute warm up, a 10-minute overground training bout, a 20-minute treadmill training bout, another 10-minute overground training bout, and a 2-minute cool down. The ‘short-interval’ HIIT protocol used 30 second bursts at maximum safe speed, alternated with 30-60 second passive recovery periods, targeting an average heart rate above 60% heart rate reserve (HRR). The MAT protocol involved continuous walking, initially targeting 40% HRR and progressing up to 60% HRR. Outcomes were assessed by blinded raters at baseline and after 4, 8 and 12 weeks of training. The current study used data from all participants enrolled in the HIT-Stroke Trial. Variables used in this analysis are described below.

### Measurement of walking capacity outcome changes

The *<u>6-Minute Walk Distance (6MWD)</u>*^15^ was used to assess walking capacity and was the primary outcome measure in the HIT-Stroke Trial.^4,14^ Among stroke survivors, 6MWD explains more variance in home and community ambulation than other laboratory measures.^16-18^

### Measurement of training parameters

- *<u>Training speed</u>* was measured with a stopwatch at the beginning and end of both overground training bouts, and was observed on the treadmill display during each burst in the treadmill training bout. Training speed for each session was the peak speed from each of the three bouts averaged together. Training speeds were expressed relative to each participant’s self-selected gait speed from the most recent testing session, by subtracting that value.
- *<u>Training heart rate</u>* data were collected continuously throughout each bout using Bluetooth monitors and were processed to calculate mean steady-state heart rate (excluding the first 3 minutes) for each bout as a percentage of HRR. This HRR calculation was the same one used for real-time monitoring, which subtracted resting heart rate in standing (just prior to the training session) from the peak heart rate recorded during any prior exercise testing visit. To mitigate the potential confound of beta-blocker medication on HRR calculation (and to maximize safety), medication was not withheld for testing or training.
- *<u>Training blood lactate</u>* concentration was measured using a finger stick and the Lactate Plus analyzer (Nova Biomedical, Waltham, MA, USA) immediately after the treadmill training bout in the middle session of each training week. For this analysis, the magnitude of lactate elevation above 2 mmol/L was calculated by subtracting 2 from each value and zeroing any negative results.
- *<u>Training step count</u>* for each session was recorded with a Stepwatch Activity Monitor (Modus Health, LLC, Edmonds, WA, USA) on the non-paretic ankle.

### Measurement of longitudinal adaptations in neuromotor gait function and aerobic capacity

- *<u>Fastest gait speed</u>* from the 10-meter walk test^19,20^ was used to measure neuromotor gait function changes. The short distance and duration of this test limits any cardiorespiratory challenge, making it a more specific neuromotor assessment. Two trials were averaged for analysis at each testing time point. *<u>Self-selected gait speed</u>* was also used in a sensitivity analysis described below.
- *<u>Oxygen consumption rate (VO_2_) at the ventilatory threshold</u>* was used to measure aerobic capacity changes. The ventilatory threshold is the most specific measure of aerobic capacity available to use among stroke survivors,^21^ for whom VO_2_-peak during an exercise test is often confounded by neuromotor impairment.^21-24^ Ventilatory threshold was measured from metabolic data acquired during a standardized symptom-limited treadmill graded exercise test, using the excess CO_2_, ventilatory equivalents and V-slope methods with rater agreement.^4^ Extracted VO_2_ values were 20-second averages. *<u>Time to ventilatory threshold</u>* and *<u>VO_2_-peak</u>* were also used in sensitivity analyses described below.

### Data analysis

When describing the study sample, baseline data were also expressed as a percentage of normative predicted values for self-selected gait speed,^25^ 6MWT distance,^26^ ventilatory threshold VO_2_^27,28^ and VO_2_-peak.^27^ To minimize missing data and simplify statistical modeling and interpretation, post-intervention outcome data were extracted from the furthest completed 6MWD time point (after 4, 8 or 12 weeks of training) for each participant to assess change from baseline. Training parameters were then averaged among sessions occurring prior to that 6MWD time point. To assess whether the primary results were influenced by different intervention and assessment durations between participants or groups, we also performed a sensitivity analysis using data from 8 weeks of training (i.e. a uniform duration across participants).

Participant characteristics, training parameters and outcomes were initially compared between groups with independent t-tests. Structural equation models (SEM) were then used to test the extent to which different variables mediated the net effects of HIIT vs. MAT on change in 6MWD. SEM analyses were implemented with the ‘R’^29^ package ‘lavaan’^30^ version 0.6-10, using centered variables, maximum likelihood estimation, ‘Huber-White’ robust standard errors, and intent-to-treat methods. Missing data were handled with full-information maximum likelihood.

#### Mediating effects of training parameters

A multiple-parallel-mediator SEM was used to assess and compare the mediating effects of all training parameters simultaneously (Figure 2, left panel). Baseline 6MWD was also included as a covariate because stroke survivors with greater baseline walking function are more likely to generate higher intensities and step counts during training, and also tend to have larger 6MWD gains from locomotor exercise.^3,31^ If not controlled, this could produce a confounded (non-causal) association between training parameter(s) and 6MWD gains.

Similarly, baseline Fugl-Meyer lower limb motor (FM-LL) score^32^ was included as a covariate to control for potential confounding by motor impairment. Stroke survivors with greater motor impairment (e.g. lower FM-LE score) may not be able to increase training speed with as much biomechanical efficiency.^33^ Thus, we were concerned that higher training heart rate at a given training speed for these individuals could confound the relationship between training heart rate and 6MWD gains if we did not adjust for baseline FM-LE score.

Unconstrained covariances were also modeled among the training parameter mediators to account for likely correlations among the different parameters. The SEM used 5 regressions:

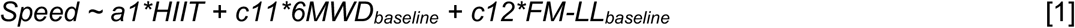

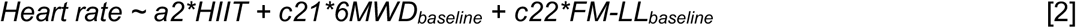

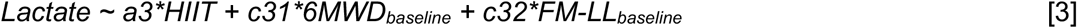

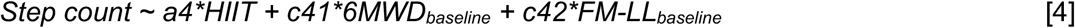

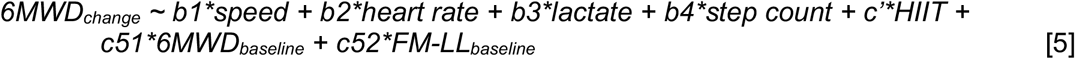

The ‘a’ path coefficient for each mediator (‘a1’ to ‘a4’) estimates the effect of HIIT vs. MAT on the mediator (after accounting for the effects of baseline 6MWD and FM-LL). The ‘b’ coefficient for each mediator (‘b1’ to ‘b4’) estimates the effect of that mediator on 6MWD change (after accounting for the effects of all the other training parameters plus treatment group, baseline 6MWD and FM-LL). The product of the corresponding ‘a’ and ‘b’ coefficients for each training parameter (e.g. ‘a4’ x ‘b4’ for step count) estimates the magnitude of the overall mediated effect of HIIT vs. MAT on 6MWD change occurring through that parameter (after accounting for the effects of the other parameters, baseline 6MWD and FM-LL).

#### Mediating effects of longitudinal adaptations in neuromotor gait function versus aerobic capacity

To assess the extent to which net 6MWT gains were mediated by net gains in neuromotor gait function versus aerobic capacity, we conducted another SEM (Figure 3, left panel) similar in structure to the previous model, but this time the potential mediators were longitudinal changes in fastest gait speed (neuromotor gait function) and ventilatory threshold VO_2_ (aerobic capacity). This SEM used 3 regressions:

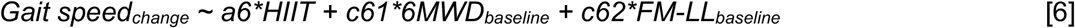

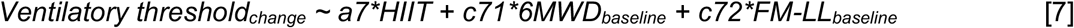

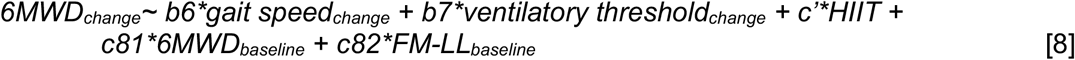

Sensitivity analyses also tested whether the results were sensitive to the specific measures selected to represent these constructs. These analyses substituted self-selected gait speed to represent neuromotor gait function or substituted time to ventilatory threshold or VO2-peak to represent aerobic capacity.

#### Model fit and multicollinearity assessments

SEM model fit was assessed with Chi-squared (χ^2^), comparative fit index (CFI), root mean square error of approximation (RMSEA) and standardized root mean square residual (SRMR), with good model fit indicated by non-significant χ^2^, χ^2^/df < 2-5, CFI > 0.90-0.95, RMSEA < 0.06-0.10 and SRMR < 0.05-0.08.^34,35^ Multicollinearity was assessed with variance inflation factors (VIFs) and condition indices, with lack of problematic multicollinearity indicated by VIFs < 5-10 and condition indices < 10-30.^36,37^

## RESULTS

Among the 55 participants randomized to HIIT (N=27) or MAT (N=28), baseline characteristics were similar between groups (Table 1). Outcome data were available from at least one post-intervention time point for 53 participants (96.4%; HIIT, 26 [96.3%]; MAT, 27 [96.4%]). The furthest completed 6MWD time point was after 4 weeks of training for 6 participants (10.9%; HIIT, 3 [11.1%]; MAT, 3 [10.7%]), after 8 weeks of training for 6 participants (10.9%; HIIT, 5 [18.5%]; MAT, 1 [3.6%]) and after the full 12-week training regimen for 41 participants (74.5%; HIIT, 18 [66.7%]; MAT, 23 [82.1%]).

**Table 1.**
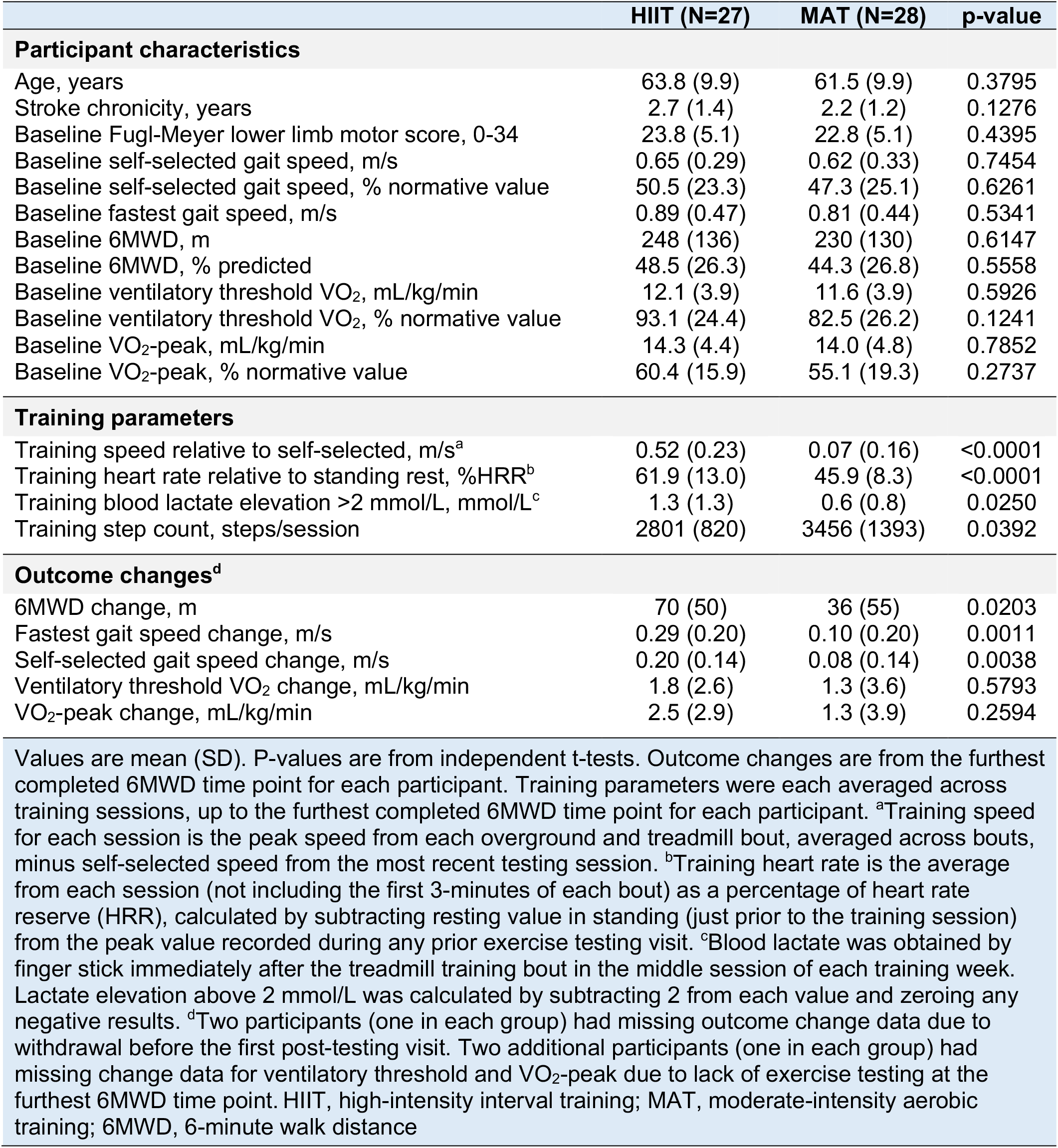
Participant characteristics, training parameters and outcomes by group.

|  |  |  |  |
| --- | --- | --- | --- |
|  | **HIIT (N=27)** | **MAT (N=28)** | **p-value** |
| **Participant characteristics** |  |  |  |
| Age, years | 63.8 (9.9) | 61.5 (9.9) | 0.3795 |
| Stroke chronicity, years | 2.7 (1.4) | 2.2 (1.2) | 0.1276 |
| Baseline Fugl-Meyer lower limb motor score, 0-34 | 23.8 (5.1) | 22.8 (5.1) | 0.4395 |
| Baseline self-selected gait speed, m/s | 0.65 (0.29) | 0.62 (0.33) | 0.7454 |
| Baseline self-selected gait speed, % normative value | 50.5 (23.3) | 47.3 (25.1) | 0.6261 |
| Baseline fastest gait speed, m/s | 0.89 (0.47) | 0.81 (0.44) | 0.5341 |
| Baseline 6MWD, m | 248 (136) | 230 (130) | 0.6147 |
| Baseline 6MWD, % predicted | 48.5 (26.3) | 44.3 (26.8) | 0.5558 |
| Baseline ventilatory threshold VO<sub>2</sub>, mL/kg/min | 12.1 (3.9) | 11.6 (3.9) | 0.5926 |
| Baseline ventilatory threshold VO<sub>2</sub>, % normative value | 93.1 (24.4) | 82.5 (26.2) | 0.1241 |
| Baseline VO<sub>2</sub>-peak, mL/kg/min | 14.3 (4.4) | 14.0 (4.8) | 0.7852 |
| Baseline VO<sub>2</sub>-peak, % normative value | 60.4 (15.9) | 55.1 (19.3) | 0.2737 |
| **Training parameters** |  |  |  |
| Training speed relative to self-selected, m/s<sup>a</sup> | 0.52 (0.23) | 0.07 (0.16) | <0.0001 |
| Training heart rate relative to standing rest, %HRR<sup>b</sup> | 61.9 (13.0) | 45.9 (8.3) | <0.0001 |
| Training blood lactate elevation >2 mmol/L, mmol/L<sup>c</sup> | 1.3 (1.3) | 0.6 (0.8) | 0.0250 |
| Training step count, steps/session | 2801 (820) | 3456 (1393) | 0.0392 |
| **Outcome changes<sup>d</sup>** |  |  |  |
| 6MWD change, m | 70 (50) | 36 (55) | 0.0203 |
| Fastest gait speed change, m/s | 0.29 (0.20) | 0.10 (0.20) | 0.0011 |
| Self-selected gait speed change, m/s | 0.20 (0.14) | 0.08 (0.14) | 0.0038 |
| Ventilatory threshold VO<sub>2</sub> change, mL/kg/min | 1.8 (2.6) | 1.3 (3.6) | 0.5793 |
| VO<sub>2</sub>-peak change, mL/kg/min | 2.5 (2.9) | 1.3 (3.9) | 0.2594 |
| Values are mean (SD). P-values are from independent t-tests. Outcome changes are from the furthest completed 6MWD time point for each participant. Training parameters were each averaged across training sessions, up to the furthest completed 6MWD time point for each participant. <sup>a</sup>Training speed for each session is the peak speed from each overground and treadmill bout, averaged across bouts, minus self-selected speed from the most recent testing session. <sup>b</sup>Training heart rate is the average from each session (not including the first 3-minutes of each bout) as a percentage of heart rate reserve (HRR), calculated by subtracting resting value in standing (just prior to the training session) from the peak value recorded during any prior exercise testing visit. <sup>c</sup>Blood lactate was obtained by finger stick immediately after the treadmill training bout in the middle session of each training week. Lactate elevation above 2 mmol/L was calculated by subtracting 2 from each value and zeroing any negative results. <sup>d</sup>Two participants (one in each group) had missing outcome change data due to withdrawal before the first post-testing visit. Two additional participants (one in each group) had missing change data for ventilatory threshold and VO<sub>2</sub>-peak due to lack of exercise testing at the furthest 6MWD time point. HIIT, high-intensity interval training; MAT, moderate-intensity aerobic training; 6MWD, 6-minute walk distance |  |  |  |

As intended, HIIT elicited significantly higher training speed, heart rate, and lactate elevation, but MAT generated higher step counts (Table 1). Changes in 6MWD, self-selected gait speed and fastest gait speed were significantly greater in HIIT vs. MAT, while changes in ventilatory threshold VO_2_ and peak VO_2_ from treadmill exercise testing were not significantly different between groups.

### Training parameters mediating net 6MWD changes induced by HIIT

HIIT-induced 6MWD improvement was found to be positively mediated by training speed and negatively mediated by training step count, with no significant mediation by training heart rate or blood lactate (Figure 2, Table 2). Training speed was a significantly greater absolute mediator than every other training parameter (p≤0.006). There was no significant residual effect of HIIT on 6MWD change after accounting for all the training parameters. In a sensitivity analysis using data from 8 weeks of training rather than each participant’s furthest completed 6MWD time point, these overall results were unchanged.

**Table 2.**
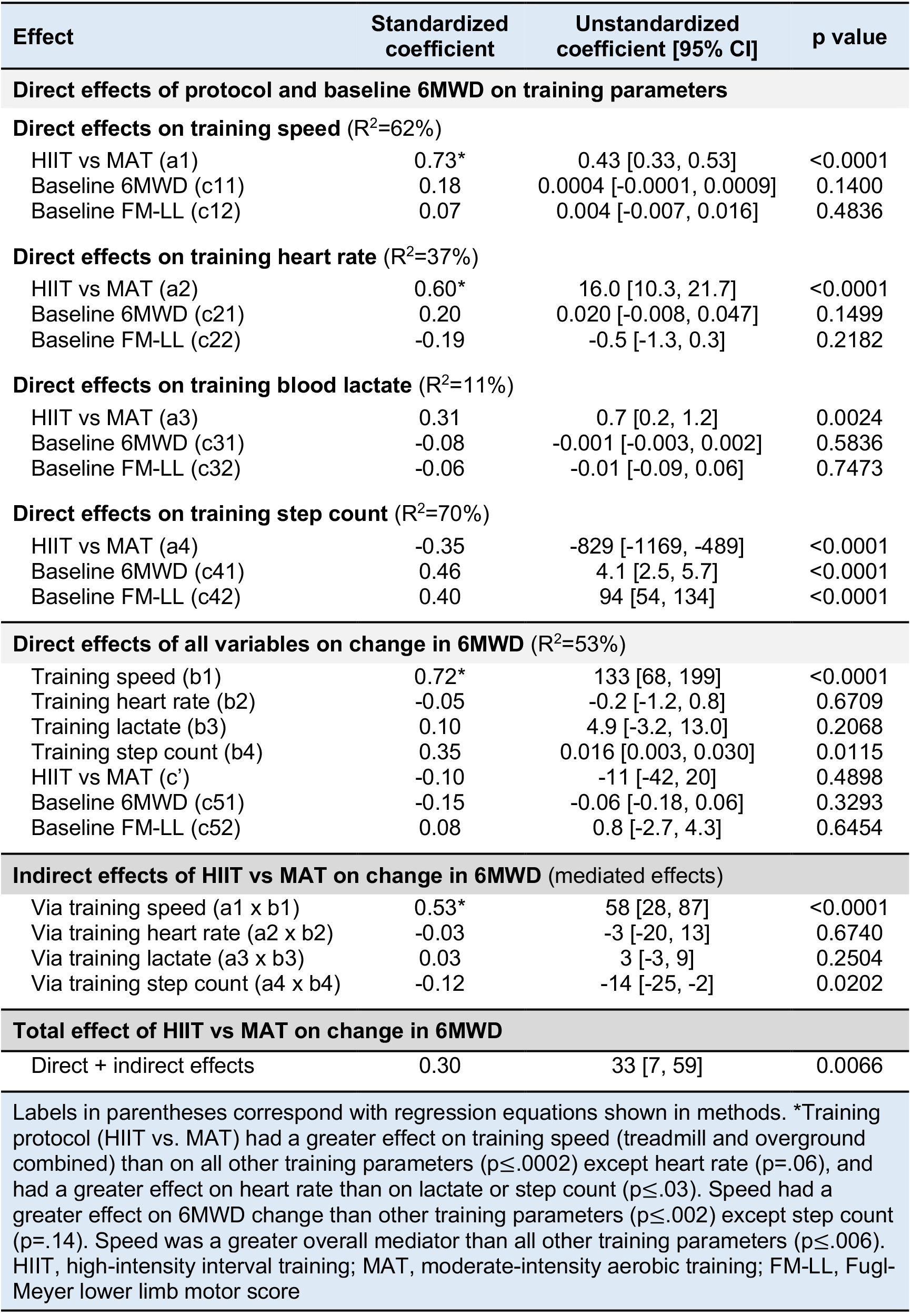
Training parameters mediating net 6-minute walk distance (6MWD) changes induced by HIIT.

Unstandardized coefficients (Table 2) describe the same associations in the original units of the variables, rather than standard deviation units. For the direct effects of HIIT vs. MAT on the training parameters, these coefficients indicated that HIIT elicited faster training speeds than MAT by 0.43 m/s [95% CI: 0.33, 0.53], higher training heart rate by 16.0% HRR [10.3, 21.7], higher blood lactate elevation by 0.7 mmol/L [0.2, 1.2] and lower training step count by 829 steps/session [-1169, -489], while controlling for baseline 6MWD.

For the direct effects of training parameters on change in 6MWD, unstandardized coefficients indicated that a 1 m/s increase in training speed (relative to self-selected speed) was associated with a 133-meter greater 6MWD gain [68, 199] (while controlling for treatment group, baseline 6MWD, baseline FM-LL and the other training parameters). In other words, each 0.1 m/s increase was associated with a 13.3-meter greater 6MWD gain [6.8, 19.9]. Likewise, each additional step taken per session was associated with a 0.016-meter greater 6MWD gain [0.003, 0.030]. Thus, each 100 step/session increase was associated with a 1.6-meter greater 6MWD gain [0.3, 3.0]. Training heart rate and blood lactate were not significantly associated with 6MWD changes.

For the overall mediated effects, unstandardized coefficients indicated that the faster training speeds with HIIT vs. MAT increased the between-group difference in 6MWD change by an estimated 58 meters [28, 87]. Likewise, the fewer steps per session with HIIT *decreased* the between-group difference in 6MWD change by an estimated 14 meters [-25, -2].

### Mediating effects of longitudinal adaptations in neuromotor gait function versus aerobic capacity on net 6MWD changes induced by HIIT

When assessing the relative contributions of neuromotor gait function changes and aerobic capacity changes to the greater 6MWD changes elicited by HIIT vs. MAT, the primary gait function variable was fastest gait speed and the primary aerobic capacity variable was ventilatory threshold VO_2_. In this model (Figure 3, Table 3), change in fastest gait speed was the only significant mediator of 6MWD changes and was a significantly greater mediator than ventilatory threshold VO_2_, with mediated effects of 37 meters [13, 61] vs. 1 meter [-3, 4] (p=0.0006). In sensitivity analyses, overall results were the same when using self-selected gait speed to represent neuromotor gait function or when using time to ventilatory threshold or VO_2_-peak to represent aerobic capacity.

**Table 3.** Mediating effects of neuromotor gait function changes vs aerobic capacity changes on net 6MWD changes induced by HIIT.

| Effect | Standardized coefficient | Unstandardized coefficient [95% CI] | p value |
| --- | --- | --- | --- |
| **Direct effects of protocol and baseline 6MWD on neuromotor vs. aerobic adaptations** |  |  |  |
| **Direct effects on fastest gait speed change ($R^2=32\%$)** |  |  |  |
| HIIT vs MAT (a6) | 0.41* | 0.18 [0.08, 0.28] | <0.0001 |
| Baseline 6MWD (c61) | 0.17 | 0.0003 [-0.0003, 0.0009] | 0.3279 |
| Baseline FM-LL (c62) | 0.20 | 0.009 [-0.007, 0.025] | 0.2958 |
| **Direct effects on ventilatory threshold change ($R^2=1\%$)** |  |  |  |
| HIIT vs MAT (a7) | 0.06 | 0.4 [-1.3, 2.1] | 0.6344 |
| Baseline 6MWD (c71) | -0.12 | -0.003 [-0.012, 0.006] | 0.5493 |
| Baseline FM-LL (c72) | 0.13 | 0.08 [-0.14, 0.30] | 0.4826 |
| **Direct effects of all variables on change in 6MWD ($R^2=75\%$)** |  |  |  |
| Fastest gait speed change (b6) | 0.81* | 204 [149, 259] | <0.0001 |
| Ventilatory threshold change (b7) | 0.12 | 2.1 [-0.9, 5.1] | 0.1561 |
| HIIT vs MAT (c') | -0.05 | -5 [-23, 12] | 0.5465 |
| Baseline 6MWD (c81) | -0.02 | -0.01 [-0.09, 0.07] | 0.8205 |
| Baseline FM-LL (c82) | 0.11 | 1.2 [-0.8, 3.2] | 0.2591 |
| **Indirect effects of HIIT vs MAT on change in 6MWD (mediated effects)** |  |  |  |
| Via gait speed change (a6 x b6) | 0.33* | 37 [13, 61] | 0.0005 |
| Via ventilatory threshold change (a7 x b7) | 0.01 | 1 [-3, 4] | 0.6422 |
| **Total effect of HIIT vs MAT on change in 6MWD** |  |  |  |
| Direct + indirect effects | 0.29 | 32 [6, 58] | 0.0093 |

### Model fit and multicollinearity diagnostics

All reported models showed good fit and no problematic multicollinearity (Table 4).

**Table 4.**
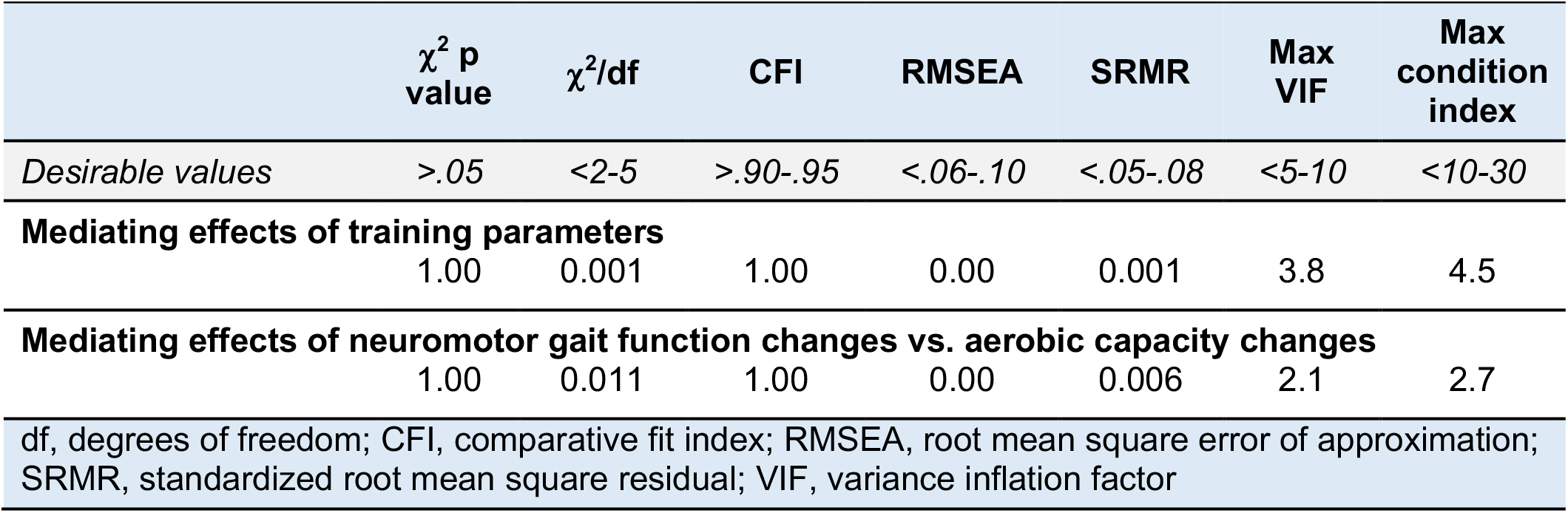
Model fit and multicollinearity measures for all models.

| Table 4. Model fit and multicollinearity measures for all models |  |  |  |  |  |  |  |
| --- | --- | --- | --- | --- | --- | --- | --- |
| | $\chi^2$ p<br>value | $\chi^2/df$ | CFI | RMSEA | SRMR | Max<br>VIF | Max<br>condition<br>index |
| *Desirable values* | *>.05* | *<2-5* | *>.90-.95* | *<.06-.10* | *<.05-.08* | *<5-10* | *<10-30* |
| **Mediating effects of training parameters** |  |  |  |  |  |  |  |
|  | 1.00 | 0.001 | 1.00 | 0.00 | 0.001 | 3.8 | 4.5 |
| **Mediating effects of neuromotor gait function changes vs. aerobic capacity changes** |  |  |  |  |  |  |  |
|  | 1.00 | 0.011 | 1.00 | 0.00 | 0.006 | 2.1 | 2.7 |
| df, degrees of freedom; CFI, comparative fit index; RMSEA, root mean square error of approximation; SRMR, standardized root mean square residual; VIF, variance inflation factor |  |  |  |  |  |  |  |

## DISCUSSION

This study sought to identify training parameters (Figure 2) and longitudinal adaptations (Figure 3) that most strongly mediate walking capacity (6MWD) gains from post-stroke HIIT. Faster training speed with HIIT vs. MAT was found to be the primary mediator of better 6MWD outcomes, increasing the net gain from HIIT by an estimated 58 meters [95% CI: 28, 87], and showing significantly stronger mediation than all other training parameters. Training step count was also found to significantly mediate 6MWD outcomes, but the *smaller* number of steps taken during HIIT vs. MAT sessions *decreased* the net 6MWD gain from HIIT by an estimated 14 meters [-25, -2].

When examining longitudinal adaptations, the greater 6MWD gain with HIIT vs. MAT was found to be exclusively mediated by change in neuromotor gait function (represented by change in short-distance fast walking speed), which increased the net 6MWD gain by an estimated 37 meters [13, 61], and showed significantly stronger mediation than change in aerobic capacity (represented by change in ventilatory threshold VO_2_). Sensitivity analyses indicated that the above findings were not solely driven by our method of extracting data from each participant’s furthest completed time point or by our selection of specific variables to represent neuromotor gait function and aerobic capacity.^11,12^

While the overall mediation effects above (i.e. the ‘a x b’ effects) are specific to the HIT and MAT protocols tested in this study, the estimated effects of each mediator on 6MWD gains (i.e. the ‘b’ effects) are designed to provide more generalizable knowledge about important training parameters and longitudinal adaptations to target during post-stroke locomotor training.^11,12^ Here, 6MWD gains were found to be positively influenced by higher speed and stepping repetition during training and greater longitudinal changes in neuromotor gait function. While additional studies are needed to confirm the generalizability of these findings, they do appear to be consistent with prior correlation analyses involving different locomotor training protocols post-stroke.^38,39^

For example, greater 6MWD gains from high-intensity continuous training have been found to correlate with higher stepping rates (a possible proxy for speed) and stepping repetitions during training, but not training heart rate.^38^ Since prior research was not designed to assess mediation, these bivariate correlations did not control for treatment group, baseline walking capacity or baseline motor impairment. By controlling for these plausible confounders, the current study provides the strongest evidence to date that training speed and stepping repetition are the most critical training parameters to target for locomotor training post-stroke. Our modeling strategy also provided novel insights about the relative importance of these parameters for increasing 6MWD. While the standardized coefficients were not significantly different between training speed and stepping repetition (0.72 vs 0.35, p=.14), these point estimates imply that training speed could impact 6MWD twice as much as stepping repetition. The unstandardized coefficients suggested that every 0.1 m/s increase in training speed would confer 13.3-meters of 6MWD gain [6.8, 19.9] and every 100 step/session increase in repetition would confer 1.6-meters of 6MWD gain [0.3, 3.0], while holding other parameters constant.

These findings suggest that maximizing speed should remain the primary training target for short interval HIIT and potentially other locomotor training protocols post-stroke. The results also insinuate that this HIIT protocol could elicit even greater 6MWD gains if it was possible to increase step count without decreasing burst speed. Unfortunately, that may be challenging, since HIIT parameter adjustments that increase step count (i.e. longer bursts, shorter recovery periods, and/or more active recovery) also tend to decrease burst speed, likely due to fatigue.^1,7,8^ Increased training time may be one option worth testing. For some individuals, it might also be possible to adaptively increase burst duration or shorten recovery duration once a speed plateau is reached within a session, contingent on speed maintenance.^3^

Contrary to our initial expectations, net 6MWD gains from HIIT vs. MAT were *not* mediated by between-group differences in training heart rate (-3 meters [-20, 13]) or aerobic adaptations (1 meter [-3, 4]). While HIIT involved significantly higher mean training heart rate than MAT by an estimated 16.0% HRR [10.3, 21.7], it did not elicit significantly greater changes in ventilatory threshold VO_2_ (0.4 mL/kg/min [-1.3, 2.1]), and 6MWD gains were not related to training heart rate (-0.2 meters per 1% HRR increase [-1.2, 0.8]) or aerobic adaptations (2.1 meters per 1 mL/kg/min increase [-0.9, 5.1]). One possible explanation for these findings is that training typically generates the greatest adaptations in the physiologic systems that are the most challenged,^40^ and neuromotor gait limitations were more severe than aerobic deconditioning in the study sample. For example, the HIIT group baseline mean for 10-m self-selected gait speed was 50.5% of normative values, and for ventilatory threshold VO_2_ was 93.1% of normative values. Another possible explanation for the lack of aerobic mediation of 6MWD changes is that neuromotor gait function might indeed be the main contributor to 6MWD performance for the average stroke survivor, as suggested by previous cross-sectional studies.^41-44^

Another finding that deviated from our hypotheses was that net 6MWD gains from HIIT vs. MAT were *not* mediated by between-group differences in blood lactate elevation during training (3 meters [-3, 9]). We initially suspected that blood lactate elevation could be an important mediator of HIIT-induced 6MWD gains because lactate accumulation physiologically differentiates HIIT from MAT,^45^ drives upregulation of circulating neurotrophins that support brain plasticity (e.g. brain-derived neurotrophic factor),^46^ and can itself act to facilitate central neuroplasticity.^47,48^ However, the current findings indicate that 6MWD gains induced by post-stroke HIIT are probably not achieved through those lactate-dependent mechanisms.

## Limitations

Causal interpretation of mediation analysis results always depends on the accuracy of modelling assumptions (e.g. no residual confounding). In this case, the randomized trial design provides strong control over confounding for the effects of treatment group on the potential mediators (i.e. the ‘a’ effects), but not for the effects of the mediators on 6MWD change (i.e. the ‘b’ effects).^9,49,50^ We controlled for the most plausible confounders by including baseline walking capacity and motor impairment measures as additional covariates in all models and testing all training parameters (or both longitudinal adaptations) simultaneously. However, we cannot rule out the possibility of residual confounding by other factors that could have produced non-causal mediation estimates or obscured causal effects.

It is also possible that differences in estimated mediation effects were at least partly driven by differences in measurement error between the tested mediators. For example, blood lactate had fewer measurement time points to average than other training parameters, which could have led to relative underestimation of its mediation effects. Another limitation was that neuromotor gait adaptation was not directly measured per-se, but was instead inferred from changes in short-distance gait speed.

## Conclusions

In chronic stroke, net gains in 6MWD from short-interval HIIT vs. MAT are primarily mediated by the faster training speeds of HIIT and by longitudinal adaptations in neuromotor gait function. Greater stepping repetition during training also appears to improve 6MWD outcomes, but training step counts are lower with this HIIT protocol vs. MAT, which seems to decrease the net 6MWD gain. These results suggest that interventions aimed at improving walking capacity post-stroke should prioritize faster training speeds and greater step counts.

## Data Availability

Data analyzed in the present study are in process to be deposited into the National Institute of Child Health and Human Development (NICHD) Data and Specimen Hub (DASH) repository

